# Prevalence and risk factors for mortality related to COVID-19 in a severely affected area of Madrid, Spain

**DOI:** 10.1101/2020.05.25.20112912

**Authors:** Ramón Pérez-Tanoira, Felipe Pérez-García, Juan Romanyk, Peña Gómez-Herruz, Teresa Arroyo, Rosa González, Lourdes Lledó García, Cristina Verdú Expósito, José Sanz Moreno, Isabel Gutiérrez, Abel Uribe Mathews, Esther López Ramos, Lara Maceda Garcia, Daniel Troncoso, Juan Cuadros-González

**Affiliations:** Departamento de Microbiología Clínica, Hospital Universitario Príncipe de Asturias, Madrid, Spain; Departamento de Biomedicina y Biotecnología, Facultad de Medicina, Universidad de Alcalá de Henares, Spain; Departamento de Medicina Interna, Hospital Universitario Príncipe de Asturias, Madrid, Spain; Departamento de Hematología, Hospital Universitario Príncipe de Asturias, Madrid, Spain; Departamento de Urgencias, Hospital Universitario Príncipe de Asturias, Madrid, Spain; Unidad de cuidados intensivos, Hospital Universitario Príncipe de Asturias, Madrid, Spain; Departamento de Bioquímica, Hospital Universitario Príncipe de Asturias, Madrid, Spain; Departamento de Medicina Preventiva, Hospital Universitario Príncipe de Asturias, Madrid, Spain

## Abstract

**BACKGROUND:** The coronavius disease 2019 (COVID-19) caused by the severe acute respiratory syndrome coronavirus 2 (SARS-CoV-2) reached Spain by 31 January 2020, in April 2020, the Comunidad de Madrid suffered one of the world’s highest crude mortality rate ratios. This study aimed to detect risk factors for mortality in patients with COVID-19.

**METHODS:** Our cohort were all consecutive adult patients (≥18 years) with laboratory-confirmed COVID-19 at a secondary hospital in Madrid, March 3-16, 2020. Clinical and laboratory data came from electronic clinical records and were compared between survivors and non-survivors, with outcomes followed up until April 4. Univariable and multivariable logistic regression methods allowed us to explore risk factors associated with in-hospital death.

**FINDINGS:** The cohort comprised 562 patients with COVID-19. Clinical records were available for evaluation for 392 patients attended at the emergency department of our hospital, of whom 199 were discharged, 85 remained hospitalized and 108 died during hospitalization. Among 311 of the hospitalized patients, 34.7% died. Of the 392 patients with records, the median age was 71.5 years (50.6-80.7); 52.6% were men. 252 (64.3%) patients had a comorbidity, hypertension being the most common: 175 (44.6%), followed by other cardiovascular disease: 102 (26.0%) and diabetes: 97 (24.7%). Multivariable regression showed increasing odds of in-hospital death associated with age over 65 (odds ratio 8.32, 95% CI 3.01–22.96; p<0.001), coronary heart disease (2.76, 1.44-5.30; 0.002), and both lower lymphocyte count (0.34, 0.17–0.68; 0.002) and higher LDH (1.25, 1.05-1.50; 0.012) per 1-unit increase and per 100 units respectively.

**INTERPRETATION:** COVID-19 was associated in our hospital at the peak of the pandemic with a crude mortality ratio of 19.2% and a mortality ratio of 34.7% in admitted patients, considerably above most of the ratios described in the Chinese series. These results leave open the question as to which factors, epidemiological or intrinsically viral, apart from age and comorbidities, can explain this difference in excess mortality.

**FUNDING:** None.

## INTRODUCTION

In December, 2019, after a series of pneumonia cases emerged in Wuhan, Hubei, China (1), sequencing analysis of samples from the lower respiratory tract indicated a coronavirus: new coronavirus 2019 (2019-nCoV) (2,3). The WHO determined that this situation should be considered a public health emergency of international interest on January 30, 2020 (2). The COVID pandemic has shattered the foundations of our healthcare system in the Comunidad de Madrid, Spain, with a total of 62 817 cases and 13 409 deaths as of May 1, 2020, and one of the highest crude mortality rate ratios worldwide (20%) (4). The Spanish government declared a state of national emergency, starting on March 15 (5).

At the beginning of the pandemic, almost all the resources of our hospital were allocated to fight this disease. By March 5, Spain had reported only 261 cases and 3 deaths (6), and the first diagnoses were confirmed in our laboratory. The initial prevalence in patients who attended our emergency department was 26%, which progressively increased to values of 35-40% in a few days, which indicated a clear pattern of community transmission. and a widespread dissemination of the virus at that time. By March 23, 1 646 cases had been confirmed, and 566 patients were admitted to our hospital with a diagnosis of COVID by PCR or unaffiliated pneumonia with a clinical diagnosis of COVID. As this is the only reference hospital in this area, we have reviewed all those confirmed by PCR diagnosis in the initial period (3 to 16 March) to determine the clinical impact of the epidemic in this period of maximal incidence, just before and shortly after the population was confined in their homes.

Here, we present details of all patients admitted to the Hospital Universitario Príncipe de Asturias (Alcalá de Henares, Madrid, Spain) with laboratory-confirmed COVID-19 and a definite clinical outcome (death or discharge) as of April 4,2020. We aim to assess mortality rates and morbidity of all patients admitted to our center at the beginning of the COVID-19 epidemic and investigate the risk factors associated with an increase in mortality.

## MATERIAL AND METHODS

### Settings and study design

We conducted the study in a middle-sized center with 453 beds and a catchment population of approximately 243 000 inhabitants in Madrid, Spain. This was a retrospective study on confirmed adult cases of COVID-19, March 3-16. A confirmed case of COVID-19 was defined as a patient with at least one compatible clinical symptom (fever > 37.5 °C, cough, or dyspnea) and a positive RT-PCR result for SARS-CoV-2 in a nasopharyngeal specimen or lower respiratory tract samples (sputum or bronchial aspirate) (7-9).

### Diagnostic procedures

The diagnosis of SARS-CoV-2 infection was performed in our centre according to the protocol established by the WHO (10). Briefly, by two automatic extractors we obtained viral RNA from clinical samples: MagCore HF16 (RBC Bioscience, Taipei, Taiwan) and Hamilton Microlab Starlet (Hamilton Company, Bonaduz, Switzerland).

RNA amplification was done with two real-time PCR platforms: VIASURE SARS-CoV-2 Real Time PCR Detection Kit (Certest Biotech, Zaragoza, Spain) and Allplex 2019-nCoV assay (Seegene, Seoul, South Korea), all used according to manufacturer’s instructions.

### Inclusion and exclusion criteria

For incidence and mortality analysis, all consecutive patients with a confirmed diagnosis by RT-PCR in nasopharyngeal-, sputum-. or lower-tract respiratory samples were included, and for the risk factors analysis, all adult patients attending the emergency department of our hospital.

Clinical data from electronic clinical records included age, sex, comorbidities (hypertension, diabetes mellitus, coronary heart disease, chronic obstructive pulmonary disease (COPD), chronic kidney disease, immunosuppression), symptoms (fever, cough, dyspnea), days from illness onset to hospital admission, laboratory data (lymphocyte count, aspartate aminotransferase (AST), alanine aminotransferase (ALT), lactate dehydrogenase, D-dimer, ferritin, and interleukin-6 (IL-6).

Treatments included any of these drugs or their combinations: a) antiretroviral treatment (darunavir/cobicistat or ritonavir/lopinavir), b) chloroquine or hydroxychloroquine, c) tocilizumab or d) type 1 interferon.

### Study definitions and outcome description

Severity of infection was classified according to WHO criteria. Briefly, patient infections were classified as: a) mild disease (patients with uncomplicated disease perhaps including non-specific symptoms) b) pneumonia (adult with pneumonia but no signs of severe pneumonia and no need for supplemental oxygen), c) severe pneumonia (fever or suspected respiratory infection, plus one of the following: respiratory rate > 30 breaths/min; severe respiratory distress or SpO2 ≤ 93% on room air), d) acute respiratory distress syndrome (within 1 week of a known clinical insult or new or worsening respiratory symptoms), e) sepsis (life-threatening organ dysfunction caused by a dysregulated host response to suspected or proven infection) and f) septic shock (persistent hypotension despite volume resuscitation, requiring vasopressors to maintain MAP ≥ 65 mmHg and serum lactate level > 2 mmol/L.) (11).

Immunosuppression at the time of presentation was considered to be present when at least one of the following conditions were documented: administration of corticosteroids (prednisone [10 mg] or equivalent) or other immunosuppressive drug within the last month, and receipt of active antineoplastic treatment. Criteria for discharging patients in this hospital were normal body temperature for three days, two negative PCR results at 24-h intervals, and resolution of all clinical symptoms.

### Statistical analysis

Continuous variables were presented as median and interquartile range (IQR) and categorical variables as proportions. We used the Mann-Whitney U-test, *χ*^2^ test, or Fisher’s exact test to compare differences between survivors and non-survivors where appropriate. For these comparisons, a *p* value of 0.05 or below was considered significant. Statistical analysis was performed with SPSS v20.0 (IBM Corp., Armonk, NY, USA).

Univariable and multivariable logistic regression models served to analyze risk factors associated with in-hospital death. The logistic regression model was adjusted by the most significant variables, with results expressed as odds ratio (OR) and 95% confidence intervals (95% CI). Considering the 392 patients attending the emergency department and to avoid overfitting in the model, nine variables were chosen for multivariable analysis on the basis of previous findings and clinical constraints.

Finally, in order to estimate the prevalence of SARS-CoV-2 infection both in the municipality Alcala de Henares and in Spain, we assessed the age distribution of all patients with COVID-19 by discharge status. We adjusted the age profile of Spanish patients by the population of the municipality Alcala de Henares and of Spain. We used 2019 population estimates from the Statistics National Institute (https://www.ine.es) to calculate the relative risk (RR) of infection with COVID-19 by age group. To calculate the RR, we followed the method of Lemaitre and colleagues (12) to explore the age profile of influenza, where RR for age group is defined as

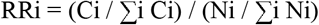

where Ci is the number of cases in age group i, and Ni is the population size of age-group i.

## Results

A total of 562 adult patients were diagnosed with SARS-CoV-2 infection in our hospital in the period of study (3-16 March, 2020), and of those, 108 died: a crude mortality rate of 19%. The mortality rate in patients attending the emergency department was 27% and for those admitted to the hospital was 34%. Considering that this hospital serves an area of 249 578 inhabitants, we determined an accumulated incidence of the disease of 225/100 000 and an accumulated mortality of 43/100 00 inhabitants.

For the risk factor analysis, all 392 patients attending the emergency department were included, but 170 other patients attending the department of occupational health or in primary care were excluded, due to a lack of key information in their medical records. Demographic and clinical findings are summarized in table 1. Briefly, their median age was 71.5 years (IQR:50.6–80.7), ranging from 23 to 99, and approximately half were male (52.6%); 311 were hospitalized, and 81 were discharged from the emergency department. Comorbidities were present in most patients, with hypertension being the most common (44.6%), followed by cardiovascular disease (26.0%) and diabetes (24.7%). These patients showed the following clinical syndromes: 106 mild disease (27.0 %), 141 pneumonia (36.0%), 116 severe pneumonia (29.6%), 27 ARDS (6.9%), and one each with sepsis and septic shock (0.3%). Of the 392, 108 (27.6%) died during hospitalization, and most patients were male (table 1).

**Table 1.**
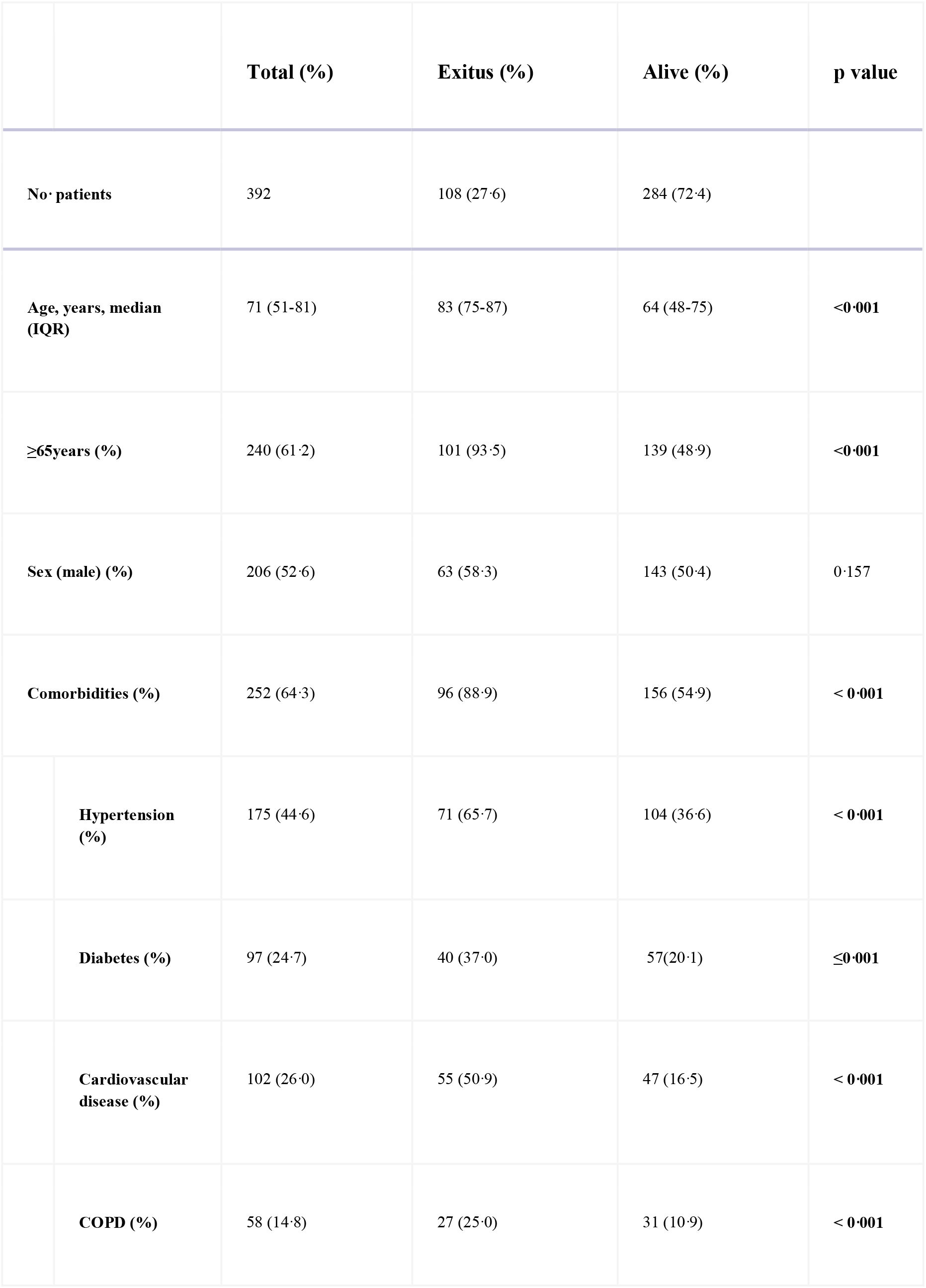
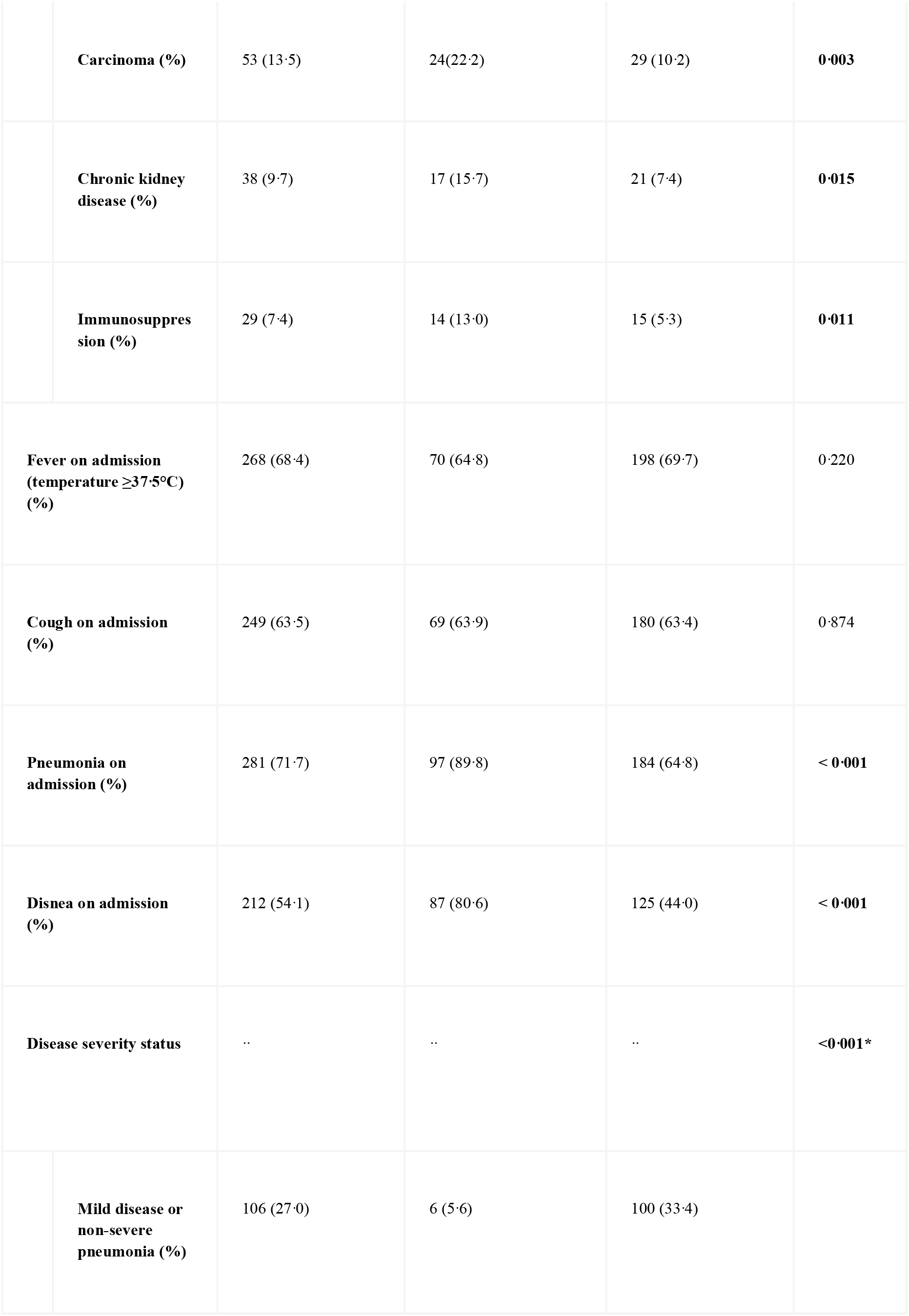
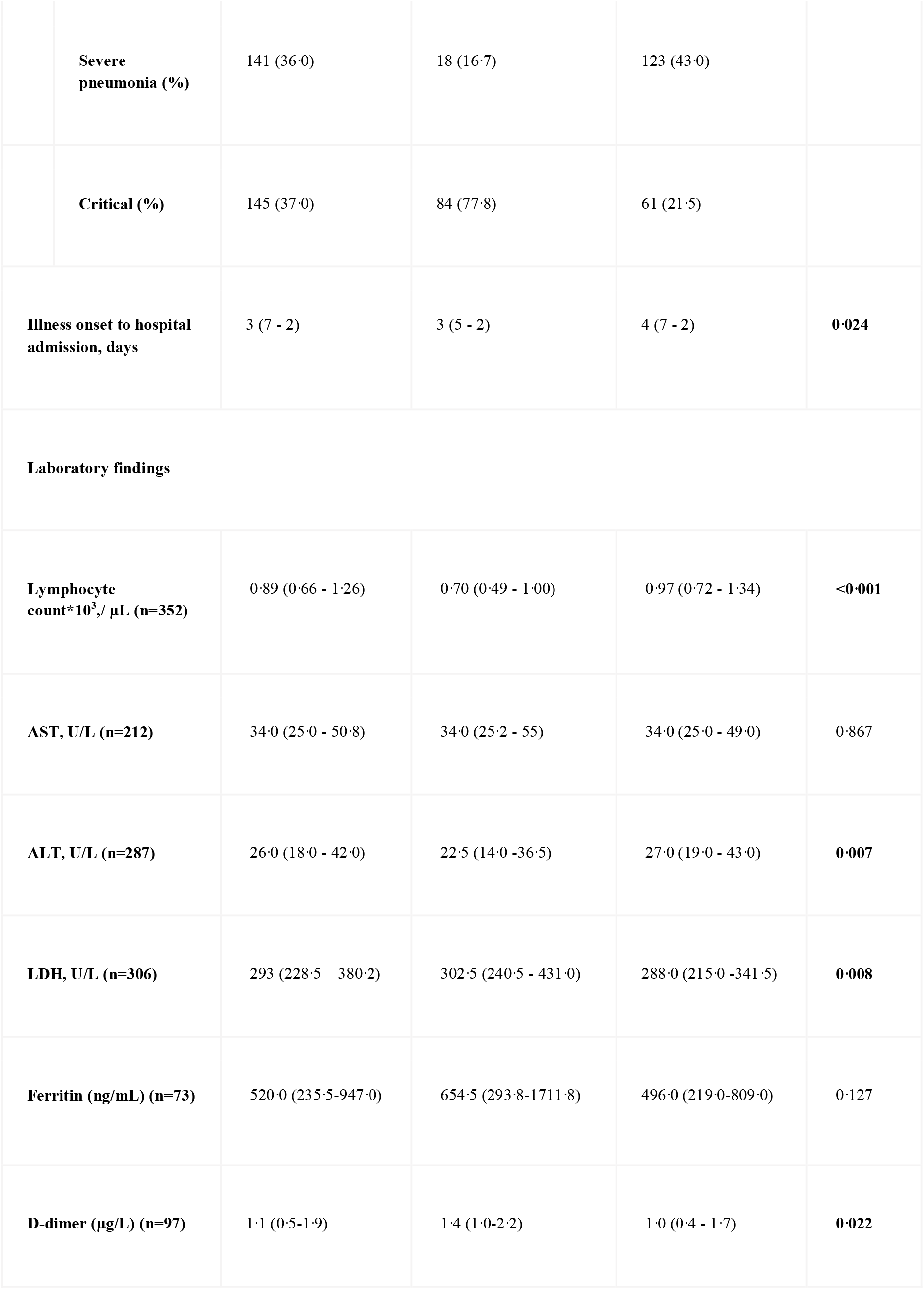
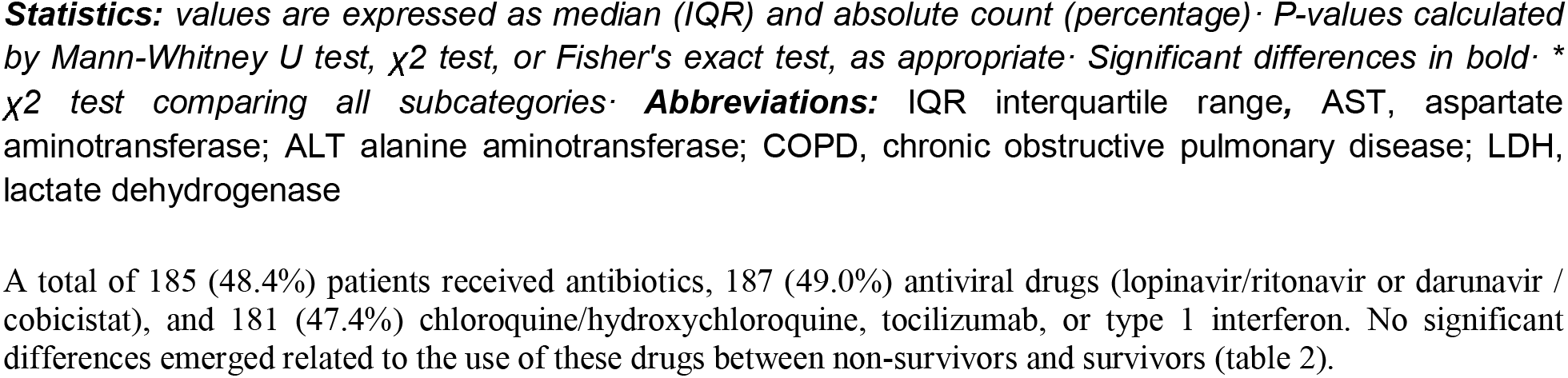
Demographic and clinical findings of patients on admission.

***Statistics:*** *values are expressed as median (IQR) and absolute count (percentage) · P-values calculated by Mann-Whitney U test, x2 test, or Fisher’s exact test, as appropriate · Significant differences in bold · * X^2^ test comparing all subcategories · **Abbreviations:*** IQR interquartile range, AST, aspartate aminotransferase; ALT alanine aminotransferase; COPD, chronic obstructive pulmonary disease; LDH, lactate dehydrogenase

A total of 185 (48.4%) patients received antibiotics, 187 (49.0%) antiviral drugs (lopinavir/ritonavir or darunavir / cobicistat), and 181 (47.4%) chloroquine/hydroxychloroquine, tocilizumab, or type 1 interferon. No significant differences emerged related to the use of these drugs between non-survivors and survivors (table 2).

**Table 2.**
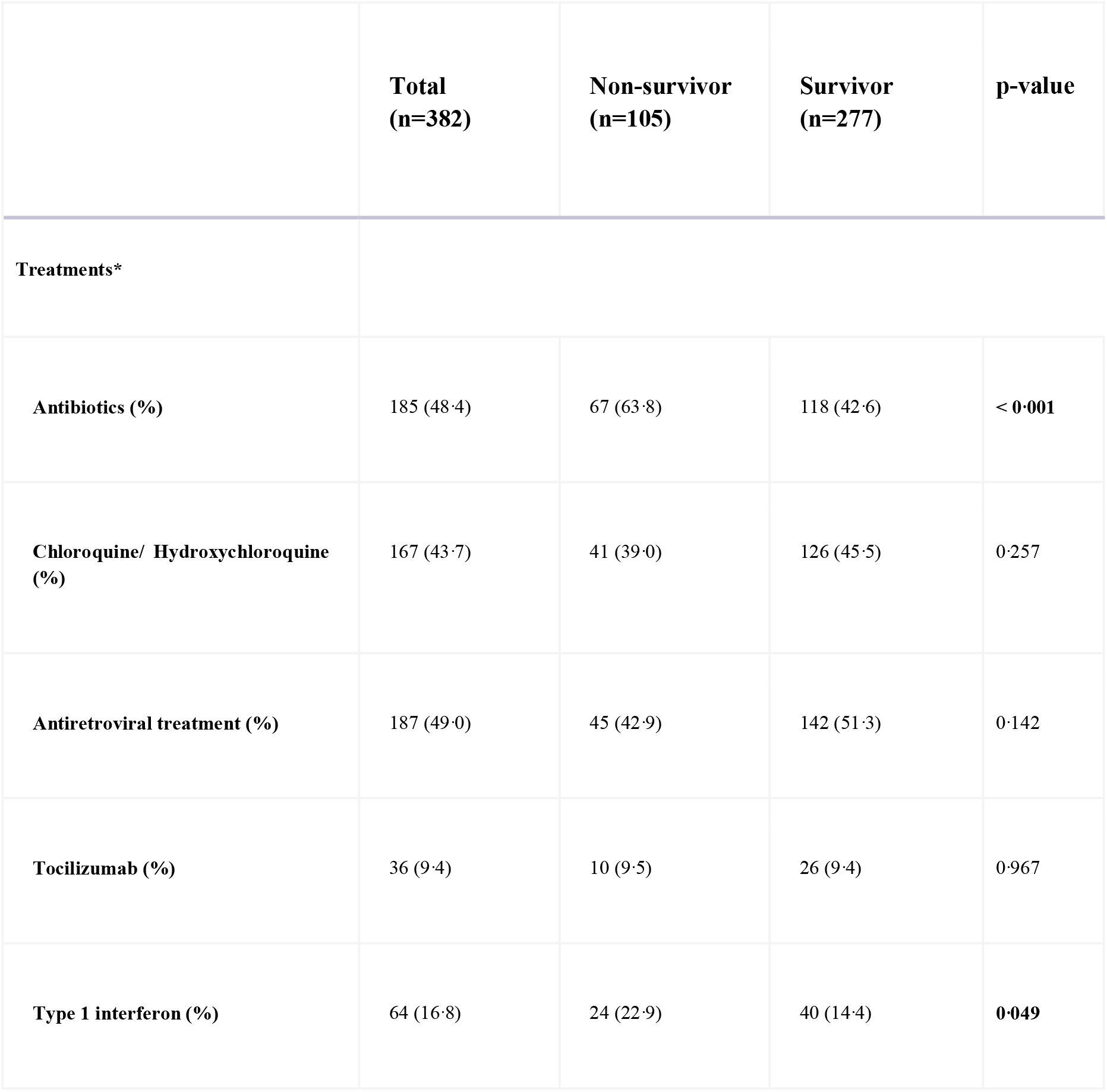
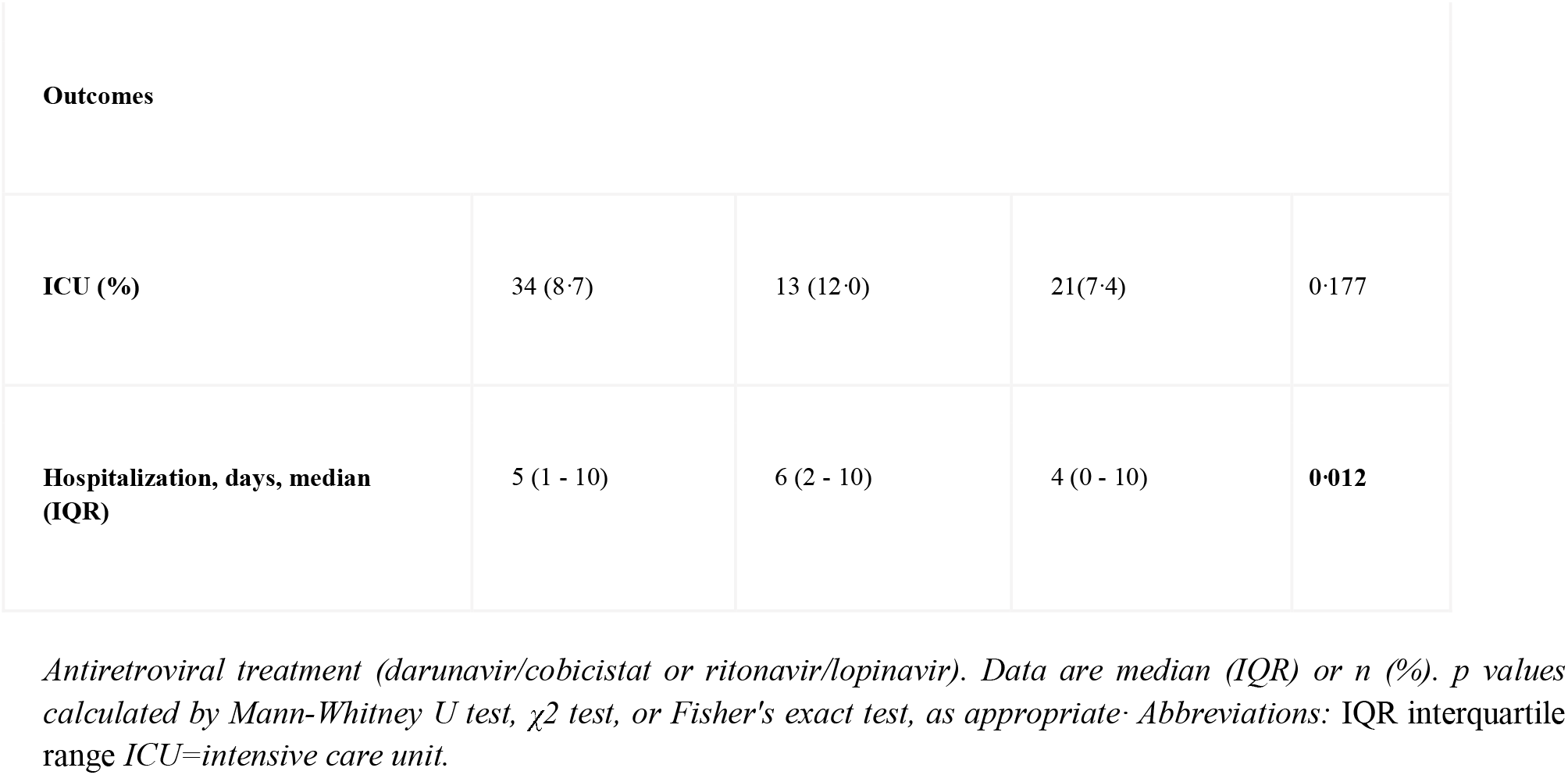
Treatments and outcomes.

|  | **Total**<br>**(n=382)** | **Non-survivor**<br>**(n=105)** | **Survivor**<br>**(n=277)** | **p-value** |
| --- | --- | --- | --- | --- |
| **Treatments\*** |  |  |  |  |
| **Antibiotics (%)** | 185 (48.4) | 67 (63.8) | 118 (42.6) | **< 0.001** |
| **Chloroquine/ Hydroxychloroquine (%)** | 167 (43.7) | 41 (39.0) | 126 (45.5) | 0.257 |
| **Antiretroviral treatment (%)** | 187 (49.0) | 45 (42.9) | 142 (51.3) | 0.142 |
| **Tocilizumab (%)** | 36 (9.4) | 10 (9.5) | 26 (9.4) | 0.967 |
| **Type 1 interferon (%)** | 64 (16.8) | 24 (22.9) | 40 (14.4) | **0.049** |

| Outcomes |  |  |  |  |
| --- | --- | --- | --- | --- |
| ICU (%) | 34 (8.7) | 13 (12.0) | 21(7.4) | 0.177 |
| Hospitalization, days, median (IQR) | 5 (1 - 10) | 6 (2 - 10) | 4 (0 - 10) | **0.012** |

*Antiretroviral treatment (darunavir/cobicistat or ritonavir/lopinavir). Data are median (IQR) or n (%). p values calculated by Mann-Whitney U test, $\chi^2$ test, or Fisher's exact test, as appropriate. Abbreviations: IQR interquartile range ICU=intensive care unit.*

In univariable analysis, odds of in-hospital death were higher in patients with any comorbidity (table 3). Age, lymphopenia, and elevated lactate dehydrogenase were also associated with death (table 3). Multivariable regression showed increasing odds of in-hospital death associated with age over 65 (odds ratio 8.32, 95% CI 3.01–22.96; p<0.001), coronary heart disease (2.76, 1.44–5.30; 0.002), and both lower lymphocyte count (0.34, 0.17–0.68; 0.002) and higher LDH (1.25, 1.05-1.50; 0.012) per 1-unit increase and per 100 units respectively (table 3).

**Table 3.**
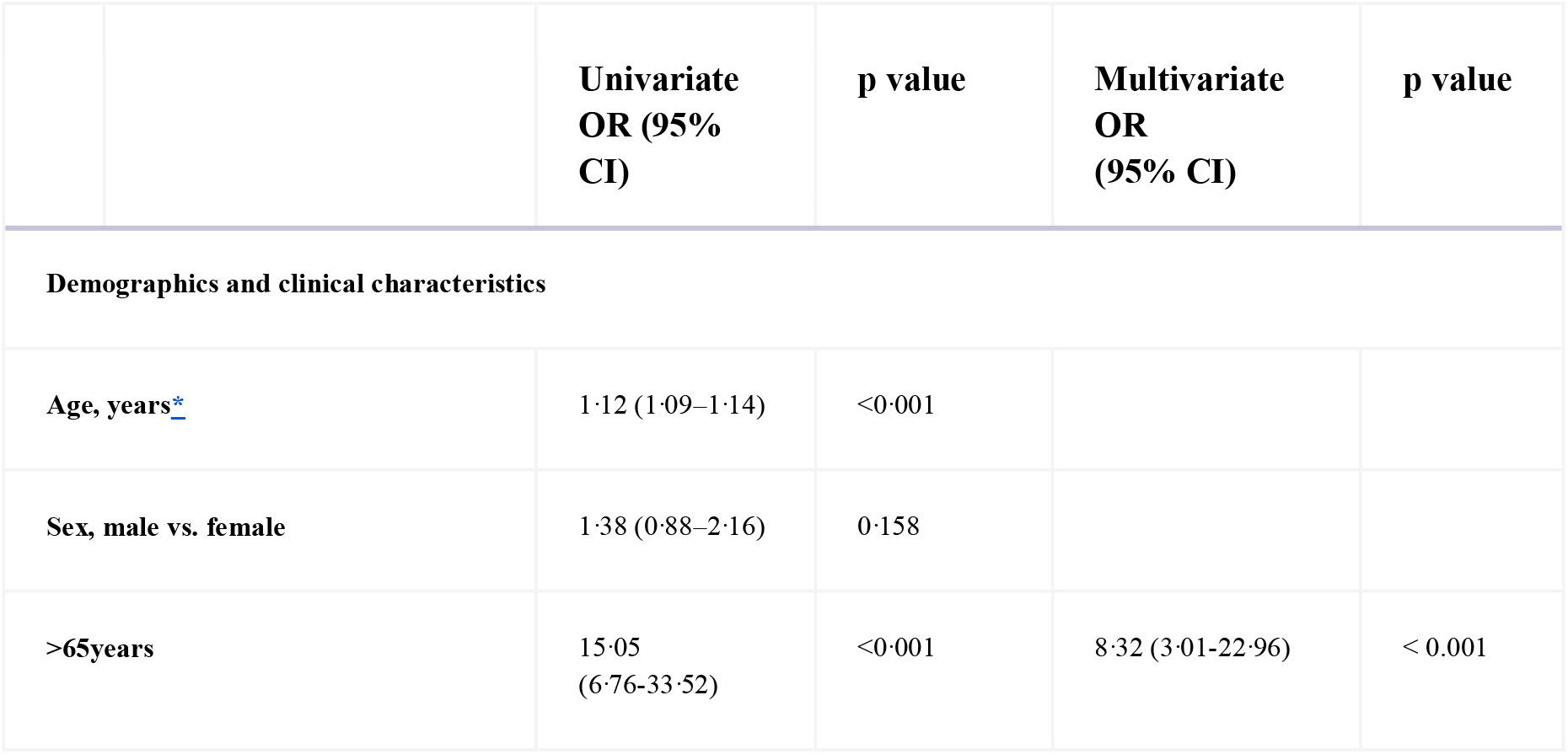
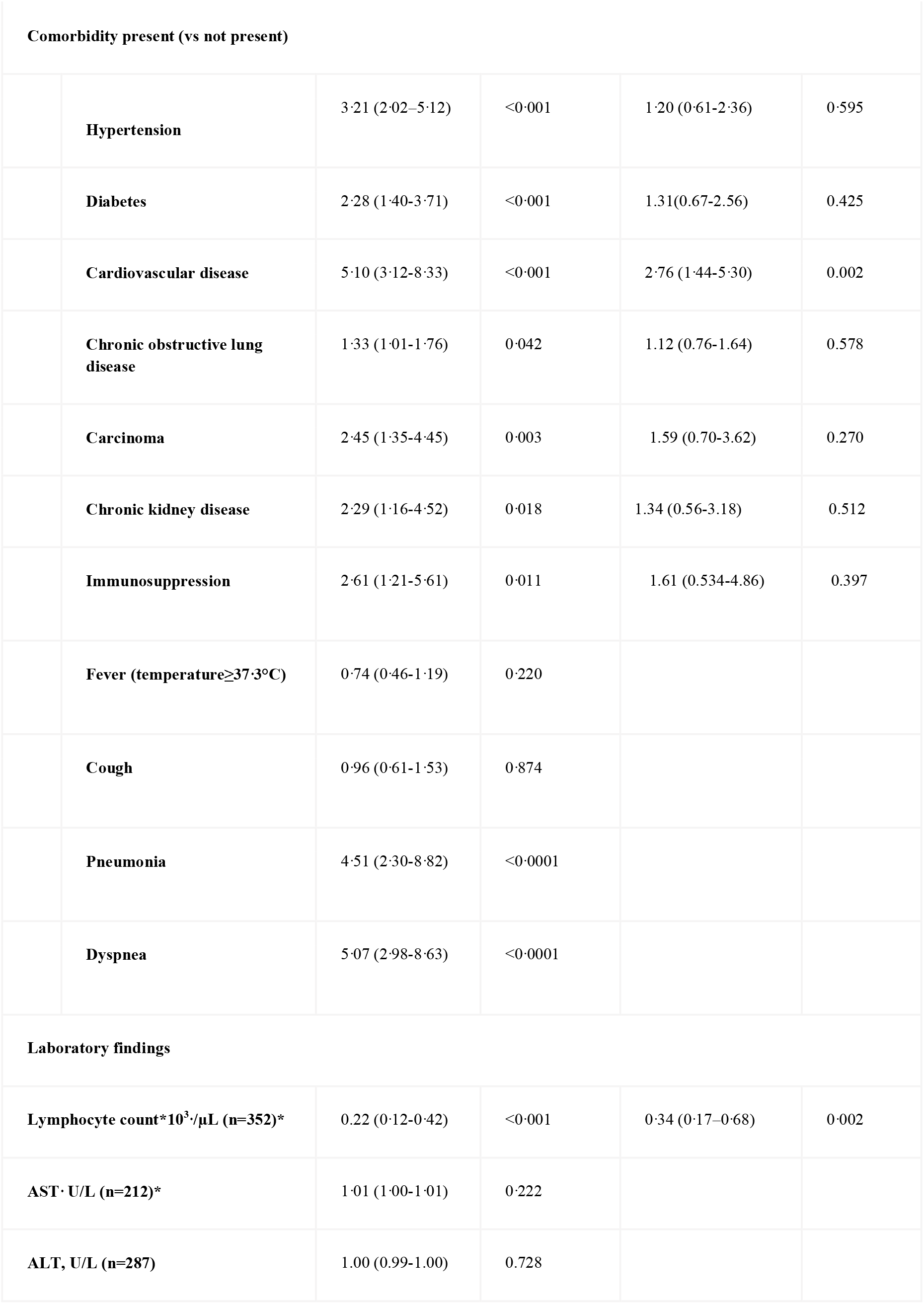
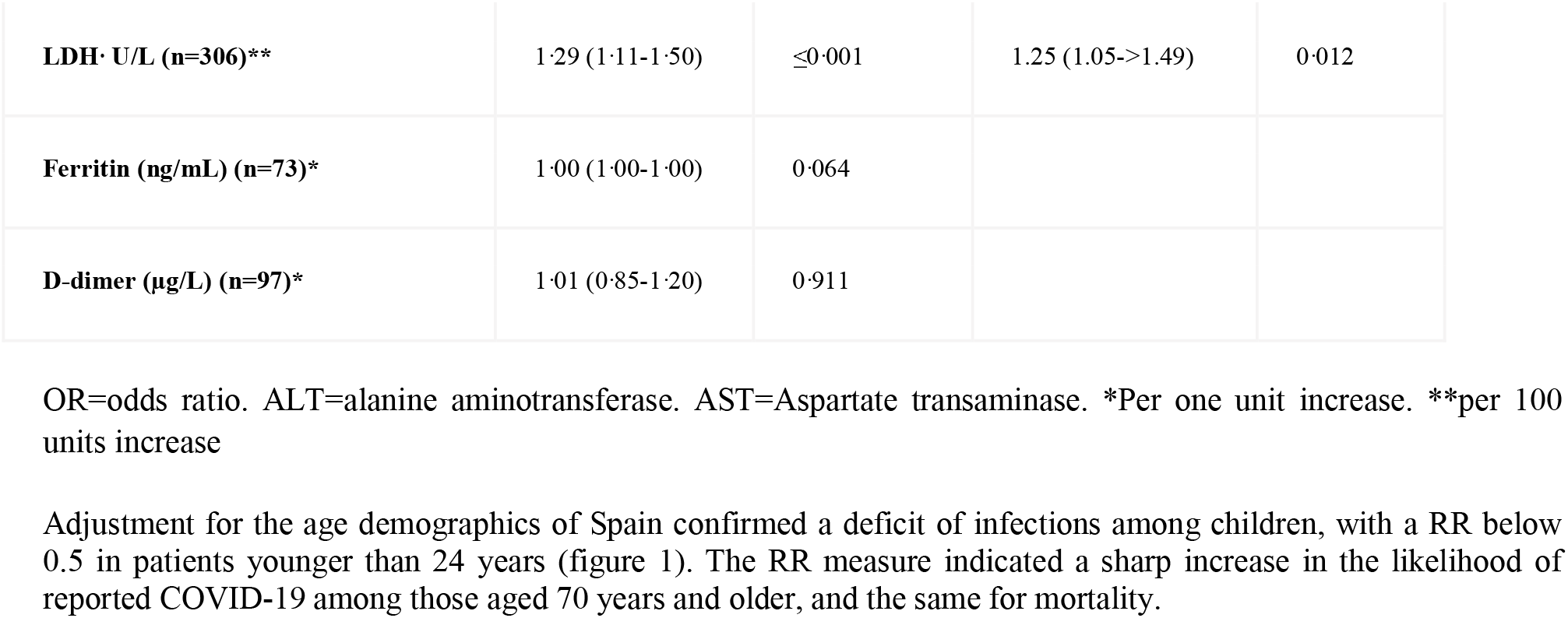
Risk factors associated with in-hospital death.

|  | Univariate<br>OR (95%<br>CI) | p value | Multivariate<br>OR<br>(95% CI) | p value |
| --- | --- | --- | --- | --- |
| Demographics and clinical characteristics |  |  |  |  |
| Age, years* | 1.12 (1.09–1.14) | <0.001 |  |  |
| Sex, male vs. female | 1.38 (0.88–2.16) | 0.158 |  |  |
| >65years | 15.05<br>(6.76-33.52) | <0.001 | 8.32 (3.01-22.96) | < 0.001 |

| Comorbidity present (vs not present) |  |  |  |  |  |
| --- | --- | --- | --- | --- | --- |
|  | **Hypertension** | 3.21 (2.02–5.12) | <0.001 | 1.20 (0.61–2.36) | 0.595 |
|  | **Diabetes** | 2.28 (1.40–3.71) | <0.001 | 1.31(0.67–2.56) | 0.425 |
|  | **Cardiovascular disease** | 5.10 (3.12–8.33) | <0.001 | 2.76 (1.44–5.30) | 0.002 |
|  | **Chronic obstructive lung disease** | 1.33 (1.01–1.76) | 0.042 | 1.12 (0.76–1.64) | 0.578 |
|  | **Carcinoma** | 2.45 (1.35–4.45) | 0.003 | 1.59 (0.70–3.62) | 0.270 |
|  | **Chronic kidney disease** | 2.29 (1.16–4.52) | 0.018 | 1.34 (0.56–3.18) | 0.512 |
|  | **Immunosuppression** | 2.61 (1.21–5.61) | 0.011 | 1.61 (0.534–4.86) | 0.397 |
|  | **Fever (temperature≥37.3°C)** | 0.74 (0.46–1.19) | 0.220 |  |  |
|  | **Cough** | 0.96 (0.61–1.53) | 0.874 |  |  |
|  | **Pneumonia** | 4.51 (2.30–8.82) | <0.0001 |  |  |
|  | **Dyspnea** | 5.07 (2.98–8.63) | <0.0001 |  |  |
| Laboratory findings |  |  |  |  |  |
|  | **Lymphocyte count\*10<sup>3</sup>/μL (n=352)\*** | 0.22 (0.12–0.42) | <0.001 | 0.34 (0.17–0.68) | 0.002 |
|  | **AST, U/L (n=212)\*** | 1.01 (1.00–1.01) | 0.222 |  |  |
|  | **ALT, U/L (n=287)** | 1.00 (0.99–1.00) | 0.728 |  |  |
| **LDH· U/L (n=306)\*\*** | 1.29 (1.11-1.50) | ≤0.001 | 1.25 (1.05->1.49) | 0.012 |
| **Ferritin (ng/mL) (n=73)\*** | 1.00 (1.00-1.00) | 0.064 |  |  |
| **D-dimer (µg/L) (n=97)\*** | 1.01 (0.85-1.20) | 0.911 |  |  |

OR=odds ratio. ALT=alanine aminotransferase. AST=Aspartate transaminase. \*Per one unit increase. \*\*per 100 units increase

Adjustment for the age demographics of Spain confirmed a deficit of infections among children, with a RR below 0.5 in patients younger than 24 years (figure 1). The RR measure indicated a sharp increase in the likelihood of reported COVID-19 among those aged 70 years and older, and the same for mortality.

**Figure 1.**
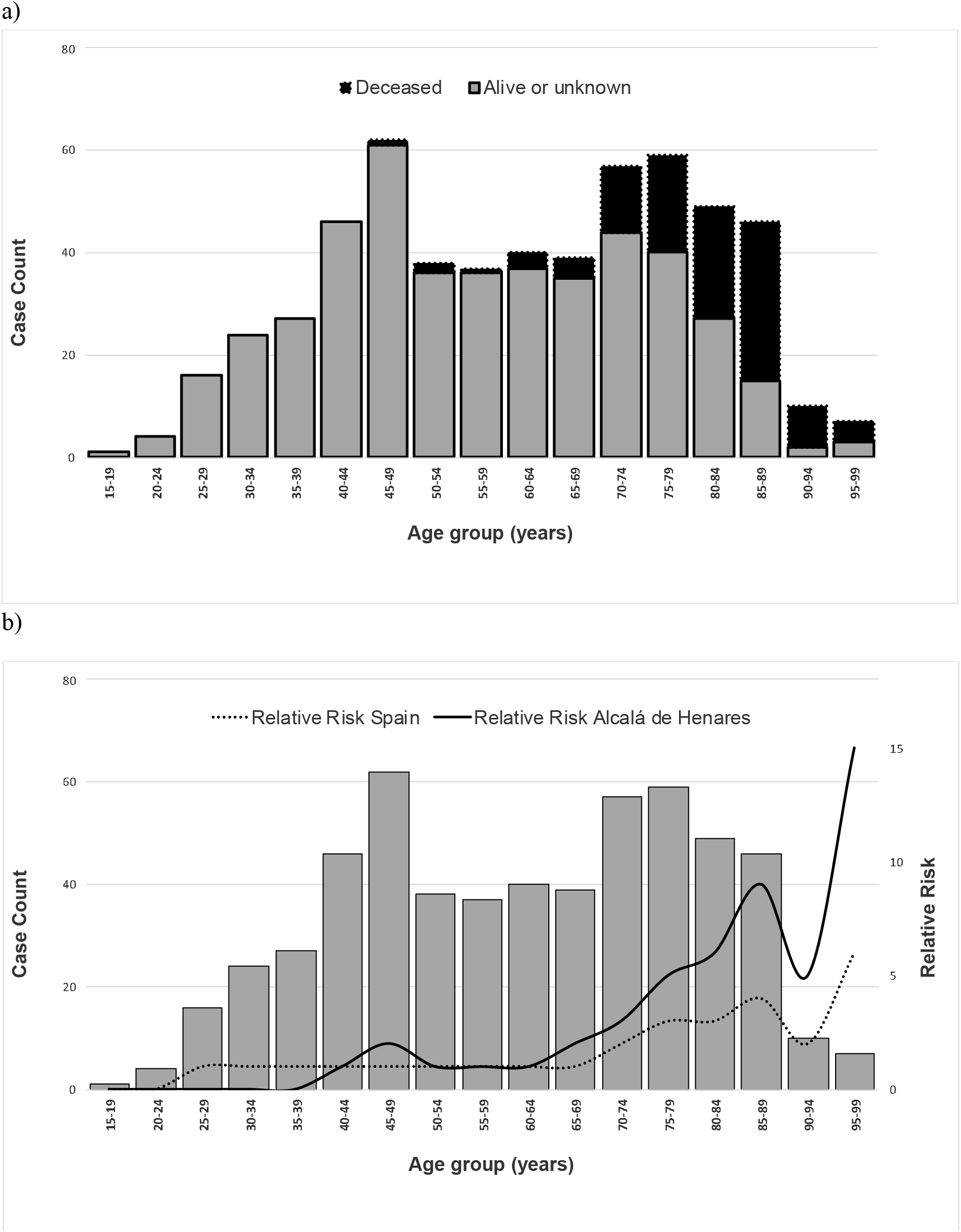
Age distribution of patients with COVID-19 from crowdsourced data (A) All 562 cases by disease outcome (alive or unknown or deceased at time of reporting); vertical bars are case counts in each age group, and dotted lines show the median age for patients who were alive or with unknown outcomes at the time of reporting and those who had died by the time of reporting. (B) Relative risk in both Spain and the municipality Alcalá de henares by 5-year age band for 562 cases reported in the Hospital Universitario Príncipe de Asturias. The data case-count quantities are bars and estimated relative risk by spline-smoothed curve. COVID-19=coronavirus disease 2019.

## Discussion

The majority of published studies regarding risk factors, severity of the disease and mortality in patients with COVID-19 come from China. The study designs range from retrospective studies, single center (13) and multicenter (8,14,15), to prospective studies (16) and meta-analysis (17). All of them mention advanced age (over aged 65) and comorbidities (especially hypertension) as factors predicting poor prognosis and mortality. Other factors mentioned are heart disease (14), diabetes (9), high lactate dehydrogenase (LDH) levels, lymphopenia or elevation of d-dimer (15,18) as well as male sex (7) or high sequential organ failure assessment score (SOFA) (14).

Few studies focus on these aspects in countries of the European Union affected by this pandemic such as Italy (19), or the United Kingdom (20), due to the short time elapsed since the start of the epidemic in Europe. In a recent multicenter study (1), the authors underline the priority of monitoring transmissibility and mortality in countries like France and Spain.

In this study, univariate analysis shows a clear statistically significative association between death and most of these risk factors: age, hypertension, diabetes, coronary heart disease, COPD, pneumonia, dyspnea, LDH levels, and lymphopenia. Multivariate logistic regression analysis reveals advanced age as a strong predictor for death from COVID-19, consistent with the findings of Du and colleagues (16). In our cohort, SARS-CoV-2 induced a community-acquired lymphopenic pneumonia characterized by lymphopenia, supporting earlier findings (21), and the existence of immunological dysregulation as an accompanying event of this critical illness.

The 27% mortality rate in patients coming through our emergency department is comparable to that of Zhou and colleagues in their reports, and is much higher than published rates in China of 11% (8) and 6% (22). This difference may depend on these series being at the beginning of their pandemic (January to February), when the disease was still poorly developed. On the other hand, 64% of our patient cohort presented with comorbidities in comparison to 51% in the cohort of Huang and colleagues which showed 15% mortality and 32% comorbidities (23).

From age 70, the relative risk of acquiring coronaviruses in our area was double that found with the same analysis but taking into account the age-structure of the Spanish general population. Our close association between advanced age and mortality rates, especially in the 70-90 age group, can be explained by our median age of 71.5, in contrast with the Chinese series with medians as low as 47 (9), 49 (24) or 56 (22).

Of our 392 patients, 85 (21.7%) were discharged and 311 (79.3%) were eventually hospitalized. The clinical spectrum in this cohort ranged from mild (27.0%) to moderate (36.0%) or severe (37.0%) infection. Nearly all admitted patients with moderate or severe infection had chest-X-ray abnormalities: of 286, 281 (98.2%), as such an abnormality was an admission criterion. No radiographic abnormalities were detectable in those with mild disease. Radiographic abnormalities are a constant in all published series of Covid-19 disease with rates ranging from 82% to 100% depending on degree of infection severity (15,23,25).

We had 185 patients (48.4%) treated with antibiotics. No differences appeared in relation to survival with any of the drugs administered to these COVID patients (lopinavir/ritonavir, chloroquine/hydroxichloroquine, tocilizumab, interferon β-1B), which are being studied in currently ongoing trials (23,24). In Spain, the health department through the AEMPS (the Spanish agency for medicines and health products) recommended a protocol for the treatment of the Covid-19, one which is available and permanently updated. However, this study was not designed for evaluation of drug efficacy, because dose and length of treatment were not registered; these results only provide background information as to the drugs used before the availability of any reliable evidence on their efficacy.

As Yang and collaborators observe, the severity of SARS-CoV-2 pneumonia places a great strain on critical care resources in hospitals, especially if their staff and resources do not match the number of patients that they must admit (26). Sometimes admission in ICUs had to be postponed or was impossible due to occupation of all the ICU beds; this can lead to deterioration of patients’ lung status and prevent their weaning, thus extending their time in the ICU. In our hospital, during the time of the study, it was necessary to increase the number of beds that normally exist almost three times, from 14 to 39 beds. Finally, the shortage of doctors and nurses is a very critical issue. Extreme decisions have been necessary, such as relocating doctors and nurses or hiring doctors who have just graduated or who have not yet been licensed.

Our research may have some limitations. Firstly, due to the retrospective study design, not all laboratory tests were done in all patients, including lactate dehydrogenase, IL-6, serum ferritin, and d-dimer. Their role may therefore be underestimated in predicting in-hospital death. An additional limitation is that some patients were transferred to another hospital, so we could not follow their outcomes. Thirdly, the number of cases may be underestimated due to frequency of collection of respiratory samples and a relatively low positive rate of detection of SARS-CoV-2 RNA in throat swabs.

## Conclusions

This study shows that the COVID epidemic had a huge impact in our community, with an accumulated incidence of the disease of 225/100 000 inhabitants and a mortality of 43/100 00 in the period 3-16 March, 2020. This difference in mortality rates in relation to previously reported series, mostly from China, leaves open the question of what other factors epidemiological or intrinsically viral, apart from age and comorbidities can explain this excess mortality. Early recognition of risk factors could be helpful in helping to quickly identify those severely affected.

## Data Availability

all data referred are availability from the corresponding author on reasonable request.

## Acknowledgements

We thank Carolyn Brimley Norris from the University of Helsinki Language Services for her aid in preparation of this manuscript.

## Compliance with Ethical Standards

### Informed consent

Since the present study is retrospective, informed consent was not required.

## Ethical approval

The study was conducted according to the ethical requirements established by the Declaration of Helsinki. The Ethics Committee of Hospital Universitario Príncipe de Asturias (Madrid) approved the study.

